# ACP risk grade: a simple mortality index for patients with confirmed or suspected severe acute respiratory syndrome coronavirus 2 disease (COVID-19) during the early stage of outbreak in Wuhan, China

**DOI:** 10.1101/2020.02.20.20025510

**Authors:** Jiatao Lu, Shufang Hu, Rong Fan, Zhihong Liu, Xueru Yin, Qiongya Wang, Qingquan Lv, Zhifang Cai, Haijun Li, Yuhai Hu, Ying Han, Hongping Hu, Wenyong Gao, Shibo Feng, Qiongfang Liu, Hui Li, Jian Sun, Jie Peng, Xuefeng Yi, Zixiao Zhou, Yabing Guo, Jinlin Hou

## Abstract

**Background:** Since the severe acute respiratory syndrome coronavirus 2 (SARS-CoV-2) disease (COVID-19) outbreaks in Wuhan, China, healthcare systems capacities in highly endemic areas have been overwhelmed. Approaches to efficient management are urgently needed and key to a quicker control of the outbreaks and casualties. We aimed to characterize the clinical features of hospitalized patients with confirmed or suspected COVID-19, and develop a mortality risk index for COVID-19 patients.

**Methods:** In this retrospective one-centre cohort study, we included all the confirmed or suspected COVID-19 patients hospitalized in a COVID-19-designated hospital from January 21 to February 5, 2020. Demographic, clinical, laboratory, radiological and clinical outcome data were collected from the hospital information system, nursing records and laboratory reports.

**Results:** Of 577 patients with at least one post-admission evaluation, the median age was 55 years (interquartile range [IQR], 39 - 66); 254 (44.0%) were men; 22.8% (100/438) were severe pneumonia on admission, and 37.7% (75/199) patients were SARS-CoV-2 positive. The clinical, laboratory and radiological data were comparable between positive and negative SARS-CoV-2 patients. During a median follow-up of 8.4 days (IQR, 5.8 - 12.0), 39 patients died with a 12-day cumulative mortality of 8.7% (95% CI, 5.9% to 11.5%). A simple mortality risk index (called ACP index), composed of Age and C-reactive Protein, was developed. By applying the ACP index, patients were categorized into three grades. The 12-day cumulative mortality in grade three (age ≥ 60 years and CRP ≥ 34 mg/L) was 33.2% (95% CI, 19.8% to 44.3%), which was significantly higher than those of grade two (age ≥ 60 years and CRP < 34 mg/L; age < 60 years and CRP ≥ 34 mg/L; 5.6% [95% CI, 0 to 11.3%]) and grade one (age < 60 years and CRP < 34 mg/L, 0%) (P <0.001), respectively.

**Conclusion:** The ACP index can predict COVID-19 related short-term mortality, which may be a useful and convenient tool for quickly establishing a COVID-19 hierarchical management system that can greatly reduce the medical burden and therefore mortality in highly endemic areas.

## Introduction

After the first case of severe acute respiratory syndrome coronavirus 2 (SARS-CoV-2) disease (COVID-19) was reported in Wuhan, China in December 2019, an increasing number of confirmed cases were identified over time worldwide.^1-5^ As of February 19, 2020, 74 576 confirmed cases with 2118 deaths were reported in China.^6^ Based on the reported clinical characteristics of patients infected with SARS-CoV-2, the overall mortality ranges from 2% to 5%, which can be even higher in the elders with severe complications.^1,7,8^ The Chinese government responded immediately and mobilized the whole country’s health resources to deal with this public health emergency. However, according to the data released at February 19, 2020, the number of confirmed cases in the city of Wuhan accounts for nearly two-thirds of the entire country while the number of deaths accounts for three-quarters of the whole country, and the mortality reached a peak of over 7% at the early stage. Wuhan city, as the epidemic area, is bearing enormous public health as well as medical burdens, and the epidemic condition of the city will robustly impact on the trends of COVID-19 epidemics nation-wide as well as worldwide. Thus, an efficient and timely triage of COVID-19 patients with high mortality risk is crucial for relieving the public health burden.

Highly notably, the current confirmation of SARS-CoV-2 infection is entirely based on the viral real-time reverse transcription polymerase chain reaction (RT-PCR) test result. At the 17th COVID-19 Prevention and Control Work Conference by the Hubei Government, it is reported that the overall positive rate of SARS-CoV-2 assay was only 30 - 40% in Wuhan among all the detected specimens from highly clinical suspected patients. This low detection rate, presumably due to a high rate of false negativity, of the current assay might lead to delay in diagnosing new COVID-19 patients as well as early discharge of infected patients who still carry the virus and thus remain as the potential infection source.

Given the current epidemiologic circumstance and the limited health system capacity, optimizing clinical management approach on COVID-19 patients is both critical and urgent. Our current study was conducted aiming to characterize the clinical features of either confirmed or suspected COVID-19 patients who were hospitalized in a COVID-19-designated hospital in Wuhan, and to develop a mortality risk index, as an evaluation tool used for establishing a COVID-19 hierarchical management system in highly endemic areas.

## Methods

### Study design and Participants

We performed a retrospective, one-centre study on patients hospitalized from January 21 to February 5, 2020, at Wuhan Hankou Hospital in Wuhan, China. Wuhan Hankou Hospital was one of the first designated by the Wuhan government to be responsible for admitting patients with confirmed or suspected COVID-19 from Wuhan in January 21, 2020. The hospital was staffed with a team of physicians from Nanfang Hospital, Southern Medical University, starting from January 24, 2020. All patients enrolled in this study were residences in Wuhan and treated according to the Chinese Guidance for COVID-19.^9^

### Data Acquisition

All data were provided by Hankou Hospital. The data collected include demographic characteristics, comorbidities, clinical signs, and symptoms, laboratory testing results, chest computed tomography (CT) scan images and clinical outcomes from the hospital information system, nursing records, and laboratory reports.

### Definition

Laboratory-confirmed case was defined as the presence of SARS-CoV-2 in respiratory specimens (including nasal and pharyngeal swabs) detected by RT-PCR. Suspected case was defined as meeting two of the following criteria: 1) fever, and/or respiratory symptoms; 2) presence of radiographic pneumonia; 3) white blood cell (WBC) counts within upper limit of normal (ULN) or hypo-lymphocytosis at early course of the disease. According to the Chinese COVID-19 prevention and control program (6th edition), severe disease was defined as meeting one of the following criteria: 1) presence of shortness of breath with a respiratory rate ≥30 breaths/minute; 2) an oxygen saturation (SpO_2_) ≤ 93% in the resting state; 3) hypoxemia defined as an arterial partial pressure of oxygen divided by the fraction of inspired oxygen (PaO2/FiO2 ratio) ≤ 300 mmHg; 4) presence of radiographic progression, defined as ≥50% increase of target lesion within 24-48 hours.^9^ The date of disease onset was defined as the day when the patients first became symptomatic. A portion of suspected patients with COVID-19 at the early stage of the outbreak did not receive SARS-CoV-2 RT-PCR testing due to the limited supplies of diagnostic detection reagents. The clinical outcomes were monitored up to February 5, 2020, the final date of follow-up.

### RT-PCR Test for SARS-CoV-2

Specimens from the upper respiratory tracts were collected from suspected patients with COVID-19 to extract SARS-CoV-2. After collection, the specimens were maintained in viral-transport medium and delivered to the local Center for Disease Control and Prevention (CDC), in Wuhan, for centralized testing. Practice of the diagnostic criteria was based on the recommendation by the National Institute for Viral Disease Control and Prevention of CDC (Available at http://ivdc.chinacdc.cn/kyjz/202001/t20200121_211337.html).

### Statistical analysis

Data were expressed as counts and percentages for categorical variables and as median (interquartile range [IQR]) for continuous variables. Qualitative and quantitative differences between subgroups were analysed using chi-square or Fisher’s exact tests for categorical parameters and the student t or Mann-Whitney’s tests for continuous parameters, as appropriate. The cumulative mortality was calculated using the Kaplan-Meier method, with the log-rank test used for comparisons. Cox proportional hazards regression analysis was used to determine risk factors of mortality. Statistical analysis was performed by Statistical Package for Social Science (SPSS version 20.0, Chicago, IL, USA) and R (Version 3.5.1). For unadjusted comparisons, a 2-side α of less than 0.05 was considered statistically significant.

The study was conducted in accordance with the guidelines of the Declaration of Helsinki and the principles of good clinical practice, and was approved by the Nanfang Hospital Ethics Committee (NFEC-2020-026). Written informed consent was waived in light of the urgent need to collect clinical data.

## Results

### Patient Characteristics at admission

A total of 577 patients with at least one post-admission evaluation were included in the final analysis. Of these studied patients, the median age was 55 years (IQR, 39 - 66) and 254 (44.0%) were men. Among 309 patients with comorbidity records, the most common chronic medical illnesses were hypertension (97 [31.4%]), diabetes mellitus (43 [13.9%]), and cardiovascular diseases (33 [10.7%]). Among all studied patients, 443 had clinical signs and symptoms documented in the medical records; the most common signs and symptoms at onset of illness were fever (323 [76.5%]), cough (258 [60.4%]), and fatigue (148 [33.4%]). The incidences of headache, sore throat, vomiting, nausea, rhinorrhea, and chest pain were less than 5%. The days from illness onset to admission were six days (IQR, 4 - 9). At admission, the most common abnormal laboratory test results were elevated C-reactive protein (CRP) (351 [80.9%]), lymphocytopenia (284 [63.1%]), and hypoalbuminemia (180 [43.7%]). The chest CT on 306 patients after admission, showed bilateral pneumonia in 291 (95.1%) patients, and involvement of ≥ three lobes in 237 (77.5%) patients. Most CT images (78.4%) showed lung opacity with high density (Table 1). Treatment mainly included antiviral therapy (486 [84.2%]), antibiotic therapy (559 [96.9%]), immunoglobulin infusion (315 [54.6%]), and use of corticosteroids (63 [10.9%]).

**Table 1.** Clinical characteristics of patients with confirmed or suspected severe acute respiratory syndrome coronavirus 2 disease.

|  | All patients<br>(N=577) | Disease severity <sup>a</sup> |  |  | SARS-CoV-2 RT-PCR test result <sup>b</sup> |  |  |
| --- | --- | --- | --- | --- | --- | --- | --- |
|  |  | Mild<br>(N=338) | Severe<br>(N=100) | P value | Positive<br>(N=75) | Negative<br>(N=124) | P value |
| **Age, years** | 55 (39 - 66) | 57 (43 - 65) | 61 (49 - 71) | 0.01 | 62 (48 - 71) | 57 (44 - 65) | 0.03 |
| ≥ 60 | 232 (40.2) | 142 (42.0) | 54 (54.0) | 0.03 | 42 (56.0) | 53 (42.7) | 0.07 |
| **Male** | 254 (44.0) | 154 (45.6) | 50 (50.0) | 0.43 | 37 (49.3) | 50 (40.3) | 0.21 |
| **Any comorbidity** | N=309 | N=212 | N=67 |  | N=46 | N=66 |  |
| Hypertension | 97 (31.4) | 59 (27.8) | 25 (37.3) | 0.14 | 20 (43.5) | 18 (27.3) | 0.08 |
| Diabetes mellitus | 43 (13.9) | 29 (13.7) | 9 (13.4) | 0.96 | 3 (6.5) | 12 (18.2) | 0.08 |
| Cardiovascular disease | 33 (10.7) | 20 (9.4) | 11 (16.4) | 0.11 | 4 (8.7) | 5 (7.6) | >0.99 |
| Chronic liver disease | 20 (6.5) | 14 (6.6) | 6 (9.0) | 0.52 | 5 (10.9) | 3 (4.5) | 0.27 |
| COPD | 6 (1.9) | 2 (0.9) | 2 (3.0) | 0.24 | 0 (0) | 0 (0) | - |
| Stoke history | 5 (1.6) | 3 (1.4) | 2 (3.0) | 0.60 | 2 (4.3) | 1 (1.5) | 0.57 |
| **Signs and symptoms** | N=443 | N=298 | N=89 |  | N=68 | N=88 |  |
| Fever | 323 (76.5) | 231 (77.5) | 58 (65.2) | 0.02 | 50 (73.5) | 71 (80.7) | 0.29 |
| Cough | 258 (60.4) | 182 (61.1) | 52 (58.4) | 0.65 | 44 (64.7) | 62 (70.5) | 0.45 |
| Fatigue | 148 (33.4) | 102 (34.2) | 25 (28.1) | 0.28 | 29 (42.6) | 33 (37.5) | 0.52 |
| Shortness of breath | 52 (10.7) | 34 (11.4) | 14 (15.7) | 0.28 | 5 (7.4) | 13 (14.8) | 0.15 |
| Muscle ache | 33 (6.6) | 18 (6.0) | 6 (6.7) | 0.81 | 2 (2.9) | 6 (6.8) | 0.47 |
| Diarrhea | 26 (6.3) | 22 (7.4) | 2 (2.2) | 0.08 | 5 (7.4) | 9 (10.2) | 0.53 |
| Headache | 39 (4.6) | 18 (6.0) | 8 (9.0) | 0.33 | 6 (8.8) | 3 (3.4) | 0.18 |
| Sore throat | 14 (3.2) | 8 (2.7) | 3 (3.4) | 0.72 | 3 (4.4) | 2 (2.3) | 0.65 |
| Vomiting | 10 (2.5) | 8 (2.7) | 2 (2.2) | >0.99 | 4 (5.9) | 1 (1.1) | 0.17 |
| Nausea | 10 (2.3) | 6 (2.0) | 0 (0) | 0.34 | 2 (2.9) | 1 (1.1) | 0.58 |
| Rhinorrhea | 6 (1.4) | 4 (1.3) | 0 (0) | 0.58 | 1 (1.5) | 0 (0) | 0.44 |
| Chest pain | 6 (1.4) | 6 (2.0) | 0 (0) | 0.34 | 0 (0) | 1 (1.1) | >0.99 |
| Days from illness onset to admission | 6 (4 - 9) | 6 (4 - 9) | 6.0 (4 - 9) | 0.72 | 6 (4 - 8) | 6 (4 - 8) | 0.09 |
| **Laboratory results** |  |  |  |  |  |  |  |
| White blood cell count, $\times 10^9/L$ | 4.7 (3.6 - 6.8) | 4.6 (3.6 - 6.6) | 5.8 (4.0 - 8.7) | 0.001 | 4.9 (3.6 - 6.8) | 5.1 (4.0 - 7.3) | 0.30 |
| >10 | 38/450 (8.4) | 17/260 (6.5) | 15/90 (16.7) | 0.004 | 5/56 (8.9) | 7/94 (7.4) | 0.76 |
| Neutrophil count, $\times 10^9/L$ | 3.3 (2.2 - 5.5) | 3.3 (2.1 - 5.2) | 5.0 (2.8 - 7.5) | <0.001 | 3.7 (2.3 - 5.4) | 4.0 (2.5 - 5.9) | 0.35 |
| >6.3 | 86/450 (19.1) | 39/260 (15.0) | 34/90 (37.8) | <0.001 | 10/56 (17.9) | 19/94 (20.2) | 0.72 |
| Lymphocyte count, $\times 10^9/L$ | 0.9 (0.6 - 1.2) | 0.9 (0.6 - 1.3) | 0.6 (0.4 - 0.9) | <0.001 | 0.9 (0.6 - 1.2) | 0.9 (0.6 - 1.2) | 0.43 |
| <1.1 | 284/450 (63.1) | 147/260 (56.5) | 77/90 (85.6) | <0.001 | 35/56 (62.5) | 58/94 (61.7) | 0.92 |
| Prothrombin time, s | 13.8 (13.0 - 14.8) | 13.9 (13.3 - 14.8) | 13.9 (13.1 - 15.2) | 0.77 | 14.2 (13.4 - 15.6) | 14.2 (13.3 - 15.1) | 0.81 |
| >16 | 39/375 (10.4) | 21/225 (9.3) | 8/82 (9.8) | 0.91 | 10/50 (20.0) | 3/68 (4.4) | 0.01 |
| D-dimer, mg/L | 0.3 (0.1 - 0.7) | 0.2 (0.1 - 0.7) | 0.5 (0.3 - 2.3) | <0.001 | 0.4 (0.1 - 1.2) | 0.3 (0.1 - 1.0) | 0.97 |
| >0.5 | 115/343 (33.5) | 66/211 (31.3) | 36/70 (51.4) | 0.002 | 21/45 (46.7) | 25/61 (41.0) | 0.56 |
| Albumin, g/L | 35.0 (31.3 - 38.2) | 35.0 (31.9 - 38.0) | 31.4 (29.2 - 35.5) | <0.001 | 33.0 (30.3 - 37.3) | 35.0 (31.0 - 37.8) | 0.47 |
| <34 | 180/412 (43.7) | 104/246 (42.3) | 58/87 (66.7) | <0.001 | 31/56 (55.4) | 34/76 (44.7) | 0.23 |
| Alanine aminotransferase, U/L | 23.0 (16.0 - 36.0) | 22.0 (16.0 - 34.0) | 28.0 (17.0 - 42.0) | 0.02 | 23.0 (17.0 - 35.0) | 26.5 (15.3 - 41.8) | 0.47 |
| >40 | 87/421 (20.7) | 46/251 (18.3) | 24/88 (27.3) | 0.07 | 11/57 (19.3) | 21/80 (26.3) | 0.34 |
| Total bilirubin, $\mu\text{mol/L}$ | 8.4 (6.0 - 11.4) | 8.4 (6.1 - 11.3) | 8.9 (6.7 - 11.3) | 0.44 | 9.2 (7.1 - 11.3) | 8.0 (6.0 - 11.0) | 0.15 |
| >20.5 | 16/421 (3.8) | 6/251 (2.4) | 5/88 (5.7) | 0.16 | 5/57 (8.8) | 2/80 (2.5) | 0.13 |
| Creatinine, $\mu\text{mol/L}$ | 63.0 (50.0 - 78.0) | 64.0 (53.0 - 79.5) | 65.5 (56.0 - 80.5) | 0.32 | 67.0 (55.0 - 83.2) | 62.0 (50.0 - 78.0) | 0.16 |
| >120 | 13/430 (3.0) | 7/252 (2.8) | 3/92 (3.3) | 0.73 | 1/54 (1.9) | 3/87 (3.4) | >0.99 |
| C-reactive protein, mg/L | 26.5 (8.2 - 36.7) | 25.7 (8.5 - 36.6) | 35.8 (25.2 - 37.9) | <0.001 | 32.6 (11.6 - 39.0) | 29.5 (7.7 - 37.0) | 0.31 |
| >5 | 351/434 (80.9) | 207/253 (81.8) | 78/84 (92.9) | 0.02 | 45/54 (83.3) | 73/91 (80.2) | 0.64 |
| **Chest CT scan** | N=306 | N=215 | N=47 |  | N=44 | N=93 |  |
| Bilateral pneumonia | 291 (95.1) | 205 (95.3) | 45 (95.7) | >0.99 | 42 (95.5) | 90 (96.8) | 0.66 |
| Number of lobes affected |  |  |  | 0.61 |  |  | 0.50 |
| 1 | 6 (2.0) | 5 (2.3) | 0 (0) |  | 1 (2.3) | 2 (2.2) |  |
| 2 | 20 (6.5) | 14 (6.5) | 2 (4.3) |  | 4 (9.1) | 3 (3.2) |  |
| ≥ 3 | 237 (77.5) | 168 (78.1) | 36 (76.6) |  | 34 (77.3) | 78 (83.9) |  |
| Not reported | 43 (14.1) | 28 (13.0) | 9 (19.1) |  | 5 (11.4) | 10 (10.8) |  |
| Lung opacity |  |  |  | 0.55 |  |  | 0.30 |
| Low density | 10 (3.3) | 8 (3.7) | 0 (0) |  | 0 (0) | 4 (4.3) |  |
| High density | 240 (78.4) | 170 (79.1) | 37 (78.7) |  | 35 (79.5) | 76 (81.7) |  |
| presence with consolidation | 3 (1.0) | 2 (0.9) | 0 (0) |  | 2 (4.5) | 1 (1.1) |  |
| Not reported | 53 (17.3) | 35 (16.3) | 10 (21.3) |  | 7 (15.9) | 12 (12.9) |  |
| **Disease severity** | N=438 |  |  |  | N=72 | N=117 | 0.20 |
| Mild | 338 (77.2) | / | / |  | 57 (79.2) | 101 (86.3) |  |
| Severe | 100 (22.8) | / | / |  | 15 (20.8) | 16 (13.7) |  |
| **SARS-CoV-2 RT-PCR test result** | N=199 | N=158 | N=31 | 0.20 |  |  |  |
| Positive | 75 (37.7) | 57 (36.1) | 15 (48.4) |  | / | / |  |
| Negative | 124 (62.3) | 101 (63.9) | 16 (51.6) |  | / | / |  |
| **Death cases** | 39 (6.8) | 11 (3.3) | 23 (23.0) | <0.001 | 1(1.3) | 0 (0) | 0.38 |

Abbreviations: RT-PCR, reverse transcription polymerase chain reaction; COPD, chronic obstructive pulmonary disease; CT, computed tomography.
Data are presented as medians (interquartile ranges, IQR) and n (%).
<sup>a</sup> The disease severity could not be evaluated in 139 patients due to lack of records of SpO<sub>2</sub>.
<sup>b</sup> The SARS-CoV-2 RT-PCR tests were conducted in 199 patients.

Among 438 patients with evaluable disease severity, 338 and 100 patients were diagnosed as mild and severe pneumonia, respectively. The cumulative mortality was significantly higher among patients with severe pneumonia than those with mild pneumonia (37.8% vs. 4.1%, P <0.001) (Figure 1). On admission, the patients with severe pneumonia, in comparison with the patients with mild pneumonia, had an older age (61 vs. 57 years, P =0.01), lower fever rate (77.5% vs. 65.2%, P =0.03), higher CRP level (35.8 vs. 25.7 mg/L, P <0.001), higher WBC count (5.8 vs. 4.6 ×10^9^/L, P =0.001), higher neutrophil count (5.0 vs. 3.3 ×10^9^/L, P <0.001), lower lymphocyte count (0.6 vs. 0.9 ×10^9^/L, P <0.001), high D-dimer (0.5 vs. 0.2 mg/L, P <0.001), lower albumin (31.4 vs. 35.0 g/L, P <0.001), and higher alanine aminotransferase (ALT) (28.0 vs. 22.0 U/L, P =0.02). There is no difference in the median days from illness onset to admission and sex, clinical signs, symptoms (except for fever), and bilateral pneumonia shown in CT scan among patients with different severities of pneumonia (Table 1).

**Figure 1.**
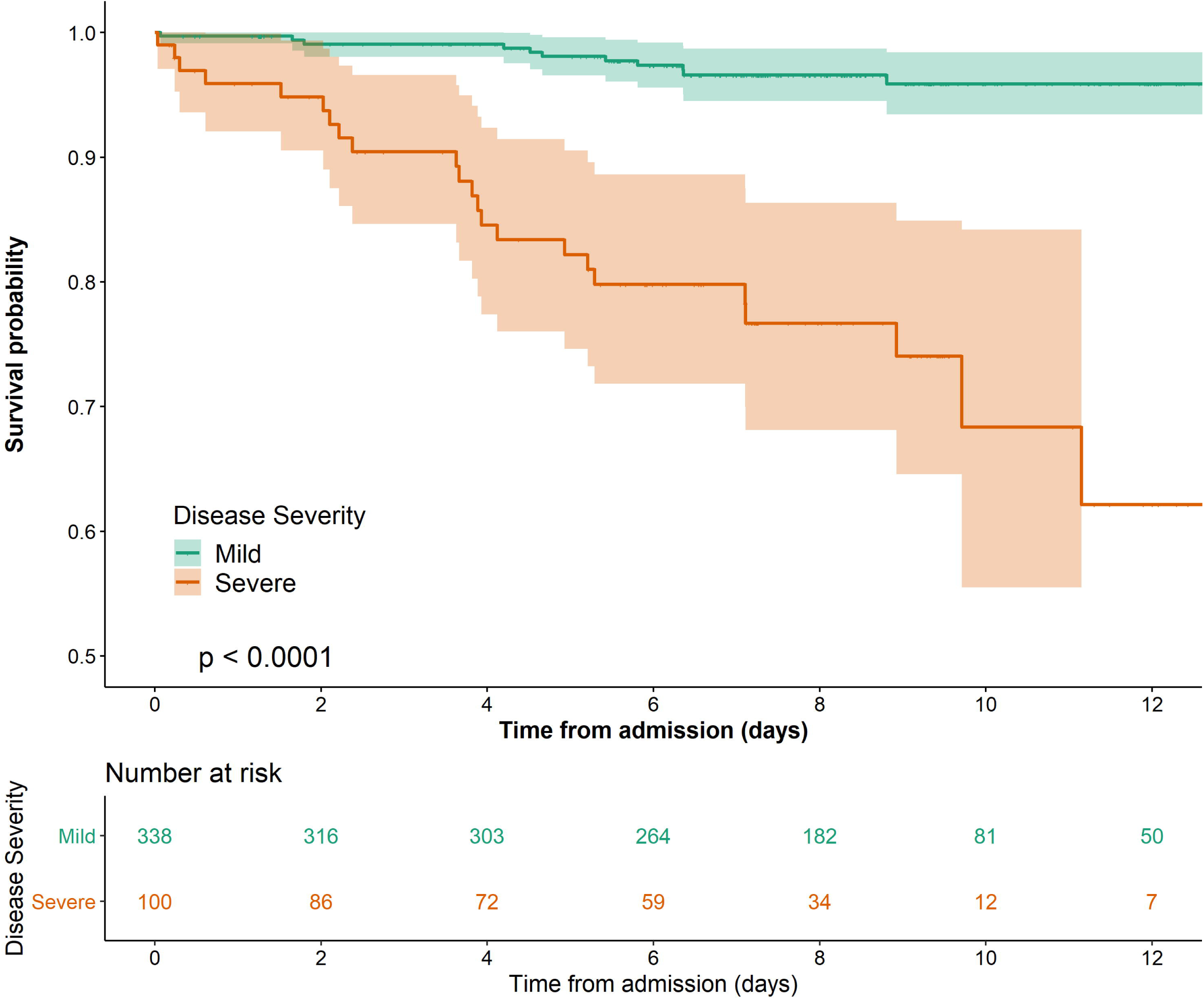
Kaplan-Meier curves of cumulative survival probability for patients with mild and severe COVID-19 within 12-day after admission. COVID-19: severe acute respiratory syndrome coronavirus 2 disease.

The SARS-CoV-2 RT-PCR tests were conducted in 199 patients. 75 (37.7%) patients had positive SARS-CoV-2 test results. Patients with positive SARS-CoV-2 test results appear to have a higher percentage of severe illness in comparison with patients who had negative SARS-CoV-2 test results (20.8% vs. 13.7%, P =0.20); however, the difference is not statistically significant. The SARS-CoV-2 test result does not seem to be related to the clinical characteristics, signs, and symptoms, as well as other laboratory test results and CT scan, except for age and chest pain (Table 1). Supplementary figure 1 showed the CT scan images of three pneumonia patients with positive (2) and negative (2) SARS-CoV-2 test results.

### Development of a COVID-19 mortality risk index

As of February 5, 2020, 39 patients died during a median follow-up of 8.4 days (IQR, 5.8 - 12.0) from admission with 12-day mortality of 8.7% (95% confidence interval [CI], 5.9% -11.5%). In the univariate cox regression analysis, age (≥ 60 vs. < 60 years), CRP (≥ 34 vs. < 34 mg/L), lymphocyte count (< 1.0 vs. ≥ 1.0 ×10^9^/L), total bilirubin (≥ 9.4 vs. < 9.4 μmol/L), creatinine (≥ 79 vs. < 79 μmol/L), albumin (< 32 vs. ≥ 32 g/L), and diabetes mellitus were associated with 12-day mortality. By multivariate cox regression analysis (with back selection), it is found that only patients’ age (hazard ratio [HR], 7.0; 95% CI, 2.1 - 23.5; P =0.002) and elevated CRP (HR, 13.5; 95% CI, 3.1 - 58.0; P <0.001) were independently significant in predicting the 12-day mortality risk (Table 2). Based upon this observation, ACP index named after the Age and C-reactive Protein was developed to stratify the mortality risk of patients with COVID-19, by which the patients were categorized into grade one (age < 60 years and CRP < 34 mg/L), grade two (age ≥ 60 years and CRP < 34 mg/L, or age < 60 years and CRP ≥ 34 mg/L) and grade three (age ≥ 60 years and CRP ≥ 34 mg/L). It was found that the ACP index is correlated to both 12-day mortality and disease severity: among 434 patients with evaluable ACP index, 194 (44.7%), 145 (33.4%), and 95 (21.9%) were assigned to grade one, grade two, and grade three, respectively. The 12-day mortality rate of grade three was 33.2% (95% CI: 19.8 - 44.3%), which was significantly higher than those of grade two (5.6% [95% CI: 0% - 11.3%]) and grade one (0%) (P <0.001), respectively (Figure 2).

**Table 2.** Risk factors associated with mortality among patients with confirmed or suspected severe acute respiratory syndrome coronavirus 2 disease.

|  | Univariate |  | Multivariate |  |
| --- | --- | --- | --- | --- |
|  | HR (95% CI) | P value | HR (95% CI) | P value |
| Age ($\geq 60$ vs. $<60$ years) | 11.2 (4.4 - 28.6) | $<0.001$ | 7.0 (2.1 - 23.5) | 0.002 |
| Sex (Male vs. Female) | 1.1 (0.6 - 2.1) | 0.71 |  |  |
| C-reactive protein ($\geq 34$ vs. $<34$ mg/L) | 25.9 (6.2 - 108.2) | $<0.001$ | 13.5 (3.1 - 58.0) | $<0.001$ |
| Lymphocyte count ($<0.5$ vs. $\geq 0.5 \times 10^9/L$) | 2.7 (1.4 - 5.5) | 0.004 | | |
| Alanine aminotransferase ($\geq 35$ vs. $<35$ U/L) | 1.8 (0.9 - 3.8) | 0.09 | | |
| Total bilirubin ($\geq 9.4$ vs. $<9.4$ $\mu\text{mol/L}$) | 2.4 (1.2 - 4.9) | 0.02 | | |
| Creatinine ($\geq 79$ vs. $<79$ $\mu\text{mol/L}$) | 2.9 (1.5 - 5.7) | 0.002 | | |
| Albumin ($<32$ vs. $\geq 32$ g/L) | 6.4 (2.9 - 13.9) | $<0.001$ | | |
| Hypertension (Yes vs. No) | 1.7 (0.9 - 3.4) | 0.11 |  |  |
| Diabetes mellitus (Yes vs. No) | 2.5 (1.2 - 5.1) | 0.02 |  |  |

Abbreviations: HR, hazard ratio; CI, confidence interval.
The threshold value of each continuous variable was the value with a maximum sum of sensitivity and specificity for predicting mortality.

**Figure 2.**
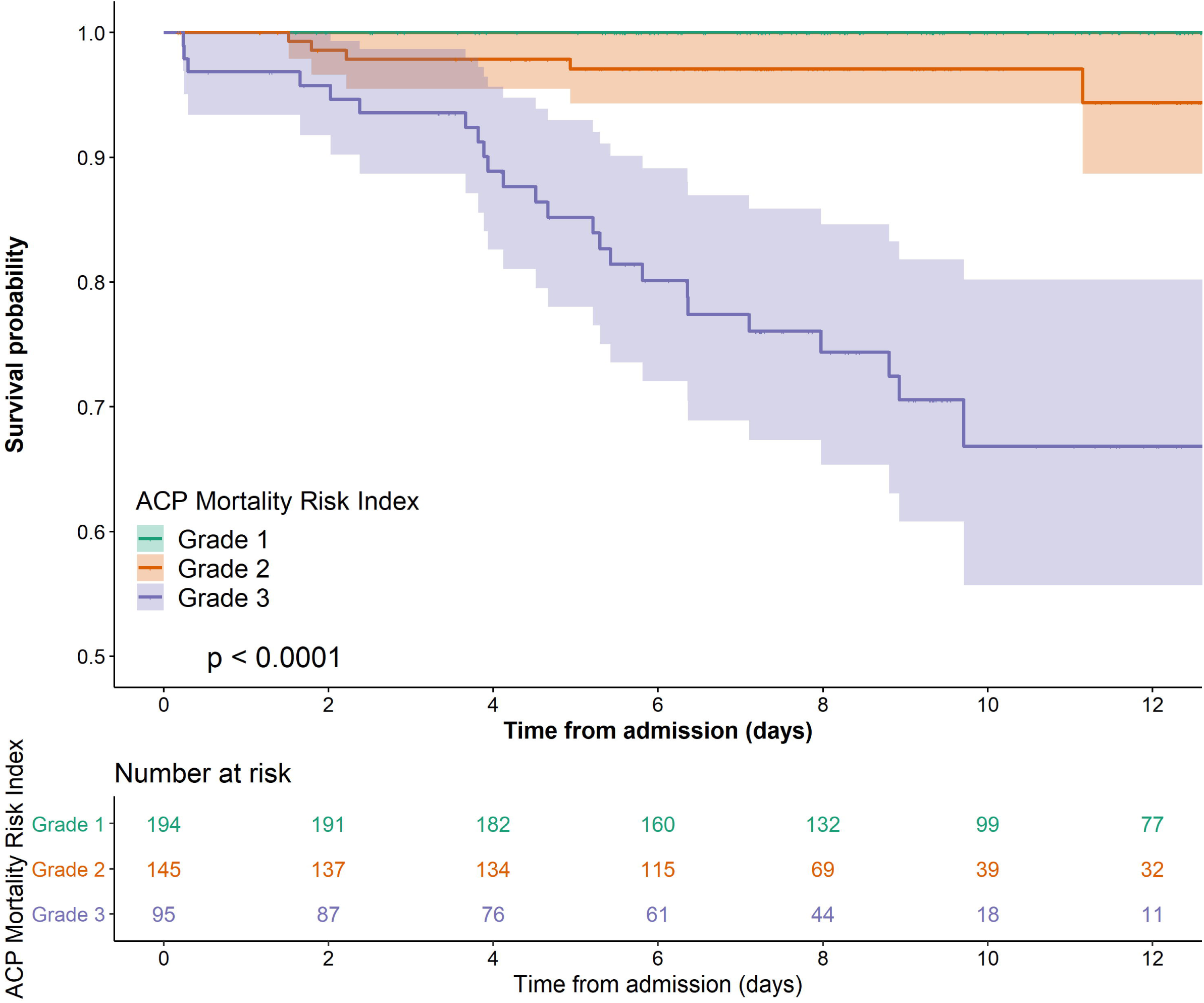
Kaplan-Meier curves of cumulative survival probability for severe acute respiratory syndrome coronavirus 2 disease patients with ACP grade one, two and three within 12-day after admission.

### Changes of laboratory test results before death

Among 12 death patients with ≥ two laboratory tests at different days, the trend of WBC, lymphocyte counts, ALT, and creatinine were evaluated. Before death, 91.7% (11/12) and 66.7% (8/12) patients experienced leukocytosis with reduction in lymphocyte counts. Among nine death patients with serial liver function tests, 44.4% (4/9) of patients had elevated ALT though most of the increases were mild (<100 U/L), except for one patient who had significantly increased ALT before death (ALT = 698 U/L). The creatinine levels remained relatively stable, except for two patients with marked elevation to 531 and 591 μmol/L on the day of death (Supplementary Figure 2). Supplementary table showed the detailed information of these 39 death patients.

## Discussion

In this study, we comprehensively described the clinical characteristics of all the patients with either confirmed or suspected COVID-19 who were hospitalized in a COVID-19 designated hospital during a period from January 21 to February 5, 2020, and then developed a simple prognostic tool called ACP index, only composed of patient’s age and CRP tested at admission, that could accurately predict the COVID-19-related mortality risk within 12 days after admission. To our knowledge, this is the first-ever study to compare the clinical characteristics of pneumonia patients who were either positive or negative for SARS-CoV-2 by RT-PCR assay, and to develop a first-ever COVID-19 mortality risk index derived from patients in highly endemic areas during early stage of outbreak.

In this large-sample size cohort, the percentage of pneumonia patients with positive results for SARS-CoV-2 result was around 40%, which was consistent with the report of Wuhan government. We found that almost all the characteristics including clinical presentations, laboratory testing results, as well as the CT scan features, were similar between pneumonia patients with or without positive results of SARS-CoV-2 RT-PCR assay. Rather than negating the diagnosis of COVID-19, the negative test results for SARS-CoV-2 are more likely to represent low sensitivity of the current RT-PCR-based assay given the similar or same clinical characteristics of the two groups of patients. Evidence highly indicated that the patients with negative results of RT-PCR assay may still have the possibility of SARS-CoV-2 infection. The poor sensitivity of the current assay could be multifactorial. The traditional RT-PCR assay has its limitations. The assay results could be influenced by specimen sources (from upper respiratory tract or lower respiratory tract) and the quality of the specimen collection. In addition, the process of virus inactivation and extraction might confer risk of viral RNA degradation in some extent and PCR inhibitors could interfere the PCR reaction, causing false-negative result.^10^ Therefore, we strongly believed that SARS-CoV-2 infections should not be ruled out on the patients with negative results of RT-PCR assay for SARS-CoV-2 when there are similar clinical presentations to COVID-19, especially in highly endemic area. Additionally, negative RT-PCR assay results for SARS-CoV-2 should not be used as a criterion to discharge seemingly improved patients after treatment from quarantine.

Considering the false-negative SARS-CoV-2 assay results, a sensitive and specific as well as fast and convenient diagnostic method needs to be developed with urgency. At present, we are conducting another promising study on the evaluation of the antibodies (immunoglobulin [Ig] G and IgM) to SARS-CoV-2 among patients in the current cohort. It is believed that the combination of detection of the pathogen and antibodies to it would improve the accuracy of SARS-CoV-2 infection diagnosis. With the increasing number of confirmed cases and deaths from SARS-CoV-2 infection, the city of Wuhan is facing a big challenge with the overwhelmed medical capacity to provide essential medical cares to all patients who need early supportive therapy and monitoring. Clinicians have to quickly triage the patients in the face of severe time and resource constraints. Thus, reliable prediction at early stage for mortality among patients with COVID-19 is valuable. The ACP index, developed on the basis of scientific evidence in this study, would provide an efficient tool in guiding decisions making on allocation of the limited medical resources to the most needed patients.

The ACP index was consisted of two components - patient’s age and CRP tested at admission. Presumably, age should be more likely to correlate with underlying medical conditions and comorbidities. In our cohort, an age of more than 60 years old indicates higher mortality in COVID-19 patients, which is consistent with previous studies on severe acute respiratory syndrome and Middle East respiratory syndrome (MERS) related pneumonia.^11,12^ Moreover, CRP, an indicator of inflammatory response, has been described as a predictor of disease progression in MERS,^13^ influenza-infected^14^ and community acquire pneumonia patients.^15^ In the current study, a CRP levels higher than 34 mg/L in the early course of COVID-19 is associated with increased mortality from SARS-CoV-2 infection, which may reflect an intense inflammatory response or/and an underlying secondary infection.

Management decision for high risk patients with COVID-19 must be made quickly, which often based upon scanty evidence. Under this pressing circumstance, the quality of evidence becomes especially crucial. The ACP index, with a high level of differentiating power, can be regarded as such quality evidence in providing an efficient, feasible and accessible approach to establish a hierarchical management system of COVID-19 in the SARS-CoV-2 highly endemic areas where medical resources are extremely limited. To be specific, it is suggested, by applying the ACP index, that patients classified in the grade one (low-risk) group, accounting for about 44.7% of the studied population, should be isolated and treated in “Mobile Cabin Hospitals”, an isolation place converted from sports stadium, convention centers, etc. with easily installed structures by the government. For patients in the grade two (medium-risk) group, which has the 12-day mortality of approximately 6%, they should be treated in the general wards, whereas the patients in the grade three group should be transferred to intensive care unit for more comprehensive treatment due to as high as 33% mortality within 12 days from admission (Figure 3). It is projected that this approach would benefit a considerable number of patients with COVID-19 by directing the medical resources appropriate for the severity of the disease, and thus reduce the mortality and save the socioeconomic resources.

**Figure 3.**
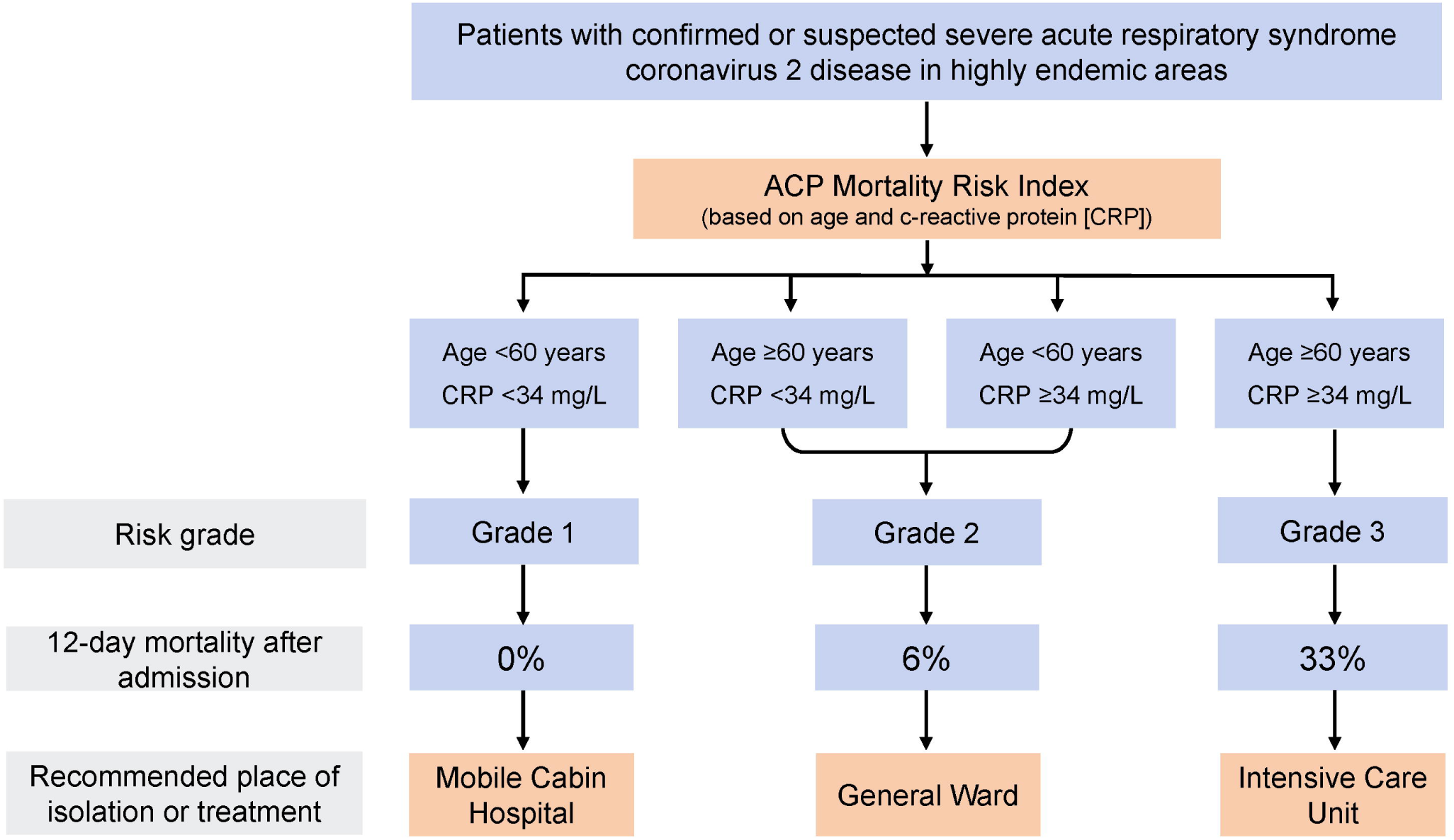
ACP index guided hierarchical management system in highly endemic areas.

We also comprehensively describe the characteristics of severe (vs. mild) and death patients in our cohort. We noticed that severe patients had high frequencies of decreased lymphocytes, higher D-dimer, and lower albumin, suggesting that severe patients may had higher probability of immune deficiency, coagulation activation, and disease exhaustion condition. In addition, we also noticed abnormal ALT and creatinine levels in most patients who died, but most of the abnormalities were mild, suggesting that COVID-19 patients may die primarily from respiratory failure caused by SARS-CoV-2 infection, rather than multiple organ failure.

The high mortality rates in our study may be due to differences in the population composition. Patients in three first designated hospitals are reasonable to have much progressed disease and higher mortality than the national-wide population, as reported by Huang et al and Chen et al.^1,8^ Notably, Hankou Hospital is the nearest medical center from the Huanan wholesale seafood market, which is reported as the first place of cluster onset. Most patients in Hankou Hospital are residents nearby and are believed to be the earliest infected population who may have close contact with live animals or the first generation of confirmed cases. In addition, Wuhan, as the endemic area, includes a considerable proportion of the elders, while those cases outside of Wuhan are mostly young people who work or study in Wuhan but return home during the Spring Festival in January 2020. These age composition differences also contribute to the prognosis quite significantly. On the other hand, suboptimal functioning of the medical care system was highly overwhelmed as too many infected subjects were presented in a very short period. Meanwhile, many patients with mild illness were home isolated in January, which lead to the reality that the current reported mortality level more precisely reflects the death risk of the severe cases.

Despite the significant findings in this report, the current study has several limitations. Firstly, some cases had incomplete documentation of the symptoms and laboratory testing given the extremely heavy work of the medical staff. Secondly, the ACP index has not been externally validated yet in an independent prospective cohort. To this end, a new prospective study is under the way to further investigate its value in triaging the COVID-19 patients. Thirdly, the patients enrolled in this study were from a tertiary-care hospital and were more likely to have underlying comorbidities and severe diseases. It is estimated that more patients would belong to the grade one category in the primary care setting. In conclusion, the ACP index can predict COVID-19 related short-term mortality, which may be a useful and convenient tool for quickly establishing a COVID-19 hierarchical management system that can greatly reduce the medical burden and therefore mortality in highly endemic areas.

## Data Availability

The data that support the findings of this study are available from the corresponding author on reasonable request.

## Author’s Contribution

Concept and design: Jinlin Hou, Yabing Guo, Jiatao Lu, Shufang Hu

Data collection: Shufang Hu, Qingquan Lv, Zhifang Cai, Haijun Li, Yuhai Hu, Ying Han, Hongping Hu,

Wenyong Gao, Shibo Feng, Qiongfang Liu, Hui Li

Data analysis: Rong Fan, Zhihong Liu, Xueru Yin

Drafting of the manuscript: Rong Fan, Zhihong Liu, Xueru Yin, Jiatao Lu, Jinlin Hou

Critical revision of the manuscript: Jian Sun, Jie Peng

Administrative, technical, or material support: Jiatao Lu, Qiongya Wang, Xuefeng Yi, Zixiao Zhou

Supervision: Jinlin Hou, Yabing Guo

## Acknowledgement

We thank all the staff of Wuhan Hankou Hospital and pay tribute to all the medical staff who are currently on the front line of the epidemic. We thank Prof. Mengfeng Li from Southern Medical University and Changcheng Zhu from New York University School of Medicine for their critical revision and professional advice on this study.

